# Circulating Tumour DNA Analysis Informing Adjuvant Chemotherapy in Stage II Colon Cancer (DYNAMIC Trial): Statistical Analysis Plan

**DOI:** 10.1101/2021.09.02.21262816

**Authors:** Jeanne Tie, Serigne N. Lo, Joshua D. Cohen, Yuxuan Wang, Rachel Wong, Jeremy Shapiro, Sam Harris, Adnan Khattak, Matthew Burge, Bert Vogelstein, Peter Gibbs

## Abstract

**Background:** Detection of circulating tumour DNA (ctDNA) after curative intent treatment reflects the presence of minimal residual disease and predicts for recurrence. DYNAMIC trial is a randomised phase II investigating the clinical utility of ctDNA-guided adjuvant treatment strategy compared with the standard of care (non-ctDNA based) approach in patients with stage II colon cancer. The primary trial endpoint is recurrence-free survival estimated from Kaplan Meier Method at 2 years. Secondary endpoints include proportion of patients treated with adjuvant chemotherapy, time-to-recurrence, overall survival, ctDNA clearance rate (investigational arm) and quality-adjusted life-years.

**Objective:** To outline and publish the pre-determined statistical analysis plan (SAP) before the database lock and the start of analysis.

**Methods:** The SAP describes basic analysis principles, methods for dealing with a range of commonly encountered data analysis issues and the specific statistical procedures for analysing efficacy and safety outcomes. The SAP was approved after closure of recruitment and before completion of patient follow-up. It outlines the planned primary analyses and a range of subgroup and sensitivity analyses regarding the clinical outcomes. Health economic outcomes are not included in this plan but will be analysed separately. The SAP will be adhered to for the final data analysis of this trial to avoid analysis bias arising from knowledge of the data.

**Trial Registration:** ANZ Clinical Trials Registry, ACTRN12615000381583. Registered on 27^th^ April 2015.

## Introduction

### Preface

Markers that better define recurrence risk for patients with stage II colon cancer are urgently required, potentially defining a subset with a high risk of recurrence that would most benefit from adjuvant chemotherapy and intensive surveillance. Markers that define a subset that clearly will not benefit from adjuvant therapy or intensive surveillance would also have major clinical utility.

Currently 20-30% of stage II patients receive adjuvant therapy, based on standard high risk clinico-pathologic criteria that only have a modest impact on recurrence risk (1, 2). Additionally, an overall survival benefit from adjuvant therapy in patients with stage II colon cancer, including those with high risk disease based on standard clinico-pathologic criteria or gene signatures, remains to be conclusively demonstrated (3-7). Rather than developing more sophisticated approaches to analysing the resected specimen to better estimate recurrence risk, which is the major focus of current efforts in stage II cancers, we are building upon the promising preliminary data from a completed observational studies demonstrating the ability of post-op circulating tumour DNA (ctDNA) analysis to detect minimal residual disease and predict for recurrence (8-10). The DYNAMIC trial will explore the clinical utility of adjuvant treatment recommendations based on a direct examination for the presence of residual disease in the patient by analysing plasma for ctDNA. In concordance with clinical trial requirements, the study was prospectively registered (ACTRN12615000381583). The trial completed the target accrual in July 2019, and the final participant will be followed up for at least 24 months.

### Purpose of the Analyses

These analyses will assess the clinical utility of ctDNA-guided adjuvant treatment strategy in comparison with the standard of care (non-ctDNA based) approach.

## Study Objectives and Endpoints

### Study Objectives

#### Primary Objective

The main purpose of this study is to demonstrate that an adjuvant therapy strategy based on ctDNA results will reduce the number of patients receiving adjuvant chemotherapy compared to standard practice without compromising the recurrence-free survival (RFS) rate after 2 years of follow-up.

#### Secondary Objectives

- To compare the time-to-recurrence (TTR) between the ctDNA-guided adjuvant approach and standard practice
- To compare the overall survival between the ctDNA-guided adjuvant approach and standard practice
- To compare the RFS/TTR between post-op ctDNA positive and negative patients in Arm A
- For Arm A (ctDNA-guided) patients with positive post-op ctDNA, to correlate end of chemotherapy ctDNA status and RFS/TTR
- To compare the RFS/TTR between post-op ctDNA positive patients in both study arms
- To compare the RFS/TTR between post-op ctDNA positive patients treated and not treated with adjuvant chemotherapy in both study arms
- To compare the RFS/TTR between post-op ctDNA negative patients who did not receive adjuvant chemotherapy in both study arms
- To correlate post-op and serial ctDNA analyses, CEA and RFS**/**TTR in ctDNA positive patients in Arm A (ctDNA-guided)
- To compare incremental costs and benefits (quality-adjusted life-years) of the two arms, i.e. comparing a ctDNA-guided strategy to the standard of care approach in adjuvant treatment decision making

#### Exploratory Objectives

- For patients with positive post-op ctDNA, to correlate post-op ctDNA levels (mutant allele fraction or other matrix) with RFS/TTR in both study arms
- For patients with positive post-op ctDNA, to correlate post-op ctDNA levels (mutant allele fraction or other matrix) with chemotherapy benefit and rate of ctDNA clearance in ctDNA-guided arm
- To correlate tumour mutation profile with post-op ctDNA detection and RFS/TTR in both study arms
- To correlate tumour infiltrating lymphocytes (TILs), post-op ctDNA and RFS/TTR in both study arms
- To assess the feasibility of measuring Fear of Cancer Recurrence levels using a validated self-reporting questionnaire in the context of a randomised adjuvant trial in both study arms

### Endpoints

#### Primary Endpoint

Recurrence-free survival rate at 2 years from randomisation. Recurrence-free survival is defined as the time from randomisation to the date of recurrence confirmation or death (from any cause). Patients who are lost to follow-up and those who are alive without recurrence will be censored at their last date of contact.

#### Key Secondary Endpoints

- Proportion of patients treated with adjuvant chemotherapy
- Time-to-recurrence (TTR) defined as the time from randomisation to the date of recurrence confirmation. Patients who died due to colon cancer but without prior diagnosis of recurrence will be coded as an event. Patients who are lost to follow-up, those who are alive without recurrence and those who died due to non-colon cancer cause will be censored.
- Overall survival (after a minimum follow-up period of 5-years) defined as time to death due to any cause. Survival time is assessed from date of randomisation to that of death.

For all time-to-event outcomes, alive patients without confirmed clinical events (recurrence or death) will be censored at their date of last contact.

#### Secondary Endpoints

additional endpoints assessed during the study will include:

- ctDNA clearance rate (investigational arm only)
- Time to ctDNA clearance (investigational arm only)
- Quality-adjusted life-years

## Study Methods

### General Study Design and Plan

This is a randomized, multi-centre, biomarker-driven adjuvant treatment study involving participants with curatively resected stage II colon cancer (Figure 1). 450 consecutive eligible participants will be enrolled at participating centres after informed consent is obtained. Participants will be randomised 2:1 to be treated according to ctDNA results (Arm A, n = 300), or per standard clinical criteria at the discretion of the treating clinician (Arm B, n = 150). Randomisation will be stratified based on the type of participating centre (metropolitan vs regional) and T stage (T3 vs T4). Patients will be consented within 21 days of surgery, with tumour samples being made available within 28 days of surgery. All participants will be scheduled for a 4-week (+/- 1 week) and 7-week (+/- 1 week) post-op blood draws for ctDNA analysis. Participants will be randomized after the week 4 blood draw. Participants who consented but failed to have a week-4 post-op blood draw will be replaced.

**Figure 1.**
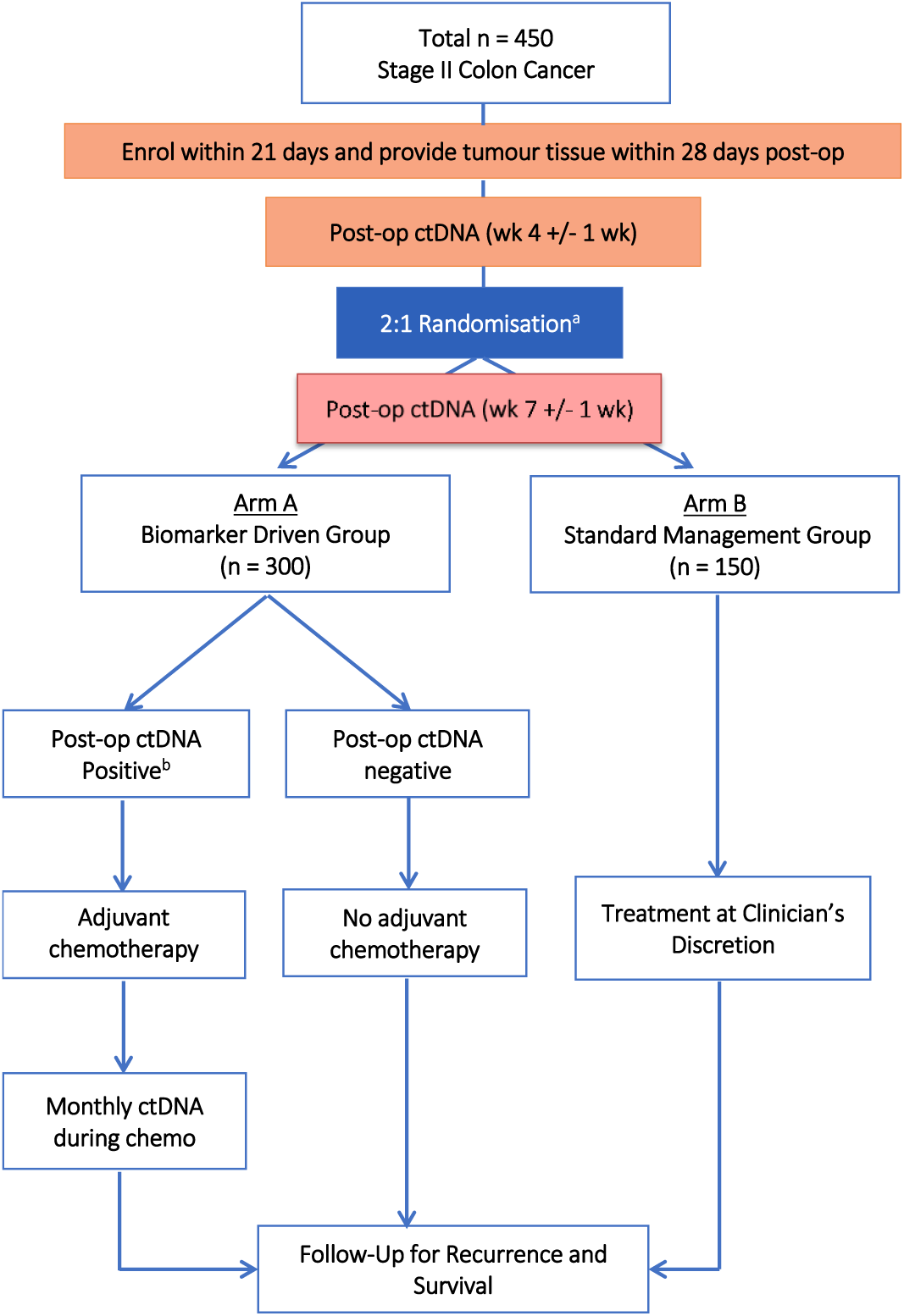
Study Design. ^a^Randomisation will occur after the first post-op ctDNA (week 4) has been collected; randomisation will be stratified by type of participating centre (metropolitan vs regional) and T stage (T3 vs T4) ^b^Patient will receive chemotherapy if they have a positive ctDNA result (either at week 4 or 7)

Both post-op samples will be analysed concurrently and ctDNA results for participants in Arm A will be made available to the treating clinician within 2-3 weeks of the second blood collection (8-10 weeks post-op). Participants /clinicians in Arm B will be blinded to their ctDNA results. In Arm A, participants with a **positive post-op ctDNA** (either at week 4 or 7) will receive 6 months of single agent fluoropyrimidine or fluoropyrimidine plus oxaliplatin adjuvant chemotherapy (clinician’s choice), and participants with confirmed **negative post-op ctDNA** (both week 4 and 7) will not be treated with adjuvant chemotherapy. Participants in Arm B will be managed at the discretion of the treating clinician. All participants will be followed for 5 years to capture recurrence and survival data.

It is anticipated that 450 eligible patients will be recruited over a 3-year period. All patients will be followed until death or study completion. It is anticipated that this study will run for approximately 8 years.

### Equivalence or Non-Inferiority Studies

The primary aim of the study is to demonstrate that patient management informed by ctDNA analysis (Arm A) leads to less chemotherapy use, (30% down to 10%), without compromising recurrence free or overall survival. While selective treatment of ctDNA positive patients in Arm A is expected to prevent disease recurrences, too few events are anticipated for this to be a statistically significant reduction.

This multi-centre randomised non-inferiority trial has been designed to determine whether patient management informed by ctDNA (arm A) is at least as efficacious as standard management (arm B) in the treatment of stage II colon cancer in terms of the primary outcome defined as 2-year RFS. Non-inferiority will be declared if the 2-year RFS rate for arm A is not worse than the 2-year RFS rate for arm B by a margin of -8.5%. The 2-year RFS in arm B is assumed to be 84% while a 2-year RFS rate of 85% is expected in Arm A. This 85% rate is computed considering 10% of the patients in Arm A will be ctDNA-positive versus 90% ctDNA-negative. The 2-year RFS rates are assumed to be 40% and 90% in ctDNA-positive and ctDNA-negative respectively.

### Inclusion-Exclusion Criteria and General Study Population

#### Inclusion Criteria

1. Participants with curatively resected stage II (T3-4, N0, M0) colon or rectal cancer
2. Patients with rectal cancer will be eligible unless they have had pre-operative combined chemotherapy and radiotherapy or are scheduled for post-operative combined chemotherapy and radiotherapy. All rectal cancer patients included in the trial must have had TME type surgery with negative (R0) resection margins.
3. A representative paraffin embedded tumour sample is available for molecular testing within 28 days after surgery. **NOTE:** Every effort should be made to submit tumour tissue sample to WEHI as soon as possible; any deviation from this timeframe will need to be discussed with the sponsor.
4. Fit for adjuvant chemotherapy
5. ECOG performance status 0-2
6. Patients that are accessible for follow-up
7. CT C/A/P within 8 weeks demonstrating no metastatic disease

#### Exclusion Criteria

1. History of another primary cancer within the last 3 years, with the exception of non-melanomatous skin cancer and carcinoma in situ of the cervix
2. Patients with multiple synchronous primary colorectal cancers
3. Patients treated with neoadjuvant chemo-radiation
4. Medical or psychiatric condition or occupational responsibilities that may preclude compliance with the protocol

### Randomisation and Blinding

Patients will be allocated to the study arms by block randomisation with block size of 3. The randomisation table was generated using R and REDCap was used to allocate each participant based on the randomisation table.

### Study Variables

|  | Baseline | 4 and 7 (+/-1) weeks post-op | Adjuvant chemo (6 months) | Follow-up visits 3 monthly for 24 months, then 6 monthly for 36 months |
| --- | --- | --- | --- | --- |
| Demographics/ ECOG | x |  |  |  |
| Serum CEA |  | x |  | x |
| Blood samples for ctDNA |  | x | (x)<sup>a</sup> |  |
| Archival tumour tissue analysis | x |  |  |  |
| Administer chemotherapy |  |  | x (if relevant) |  |
| Recurrence status |  |  |  | x<sup>b</sup> |
| Survival status |  |  |  | x |

<sup>a</sup>For Group A (ctDNA-guided) patients receiving adjuvant chemotherapy (post-op ctDNA positive patients), blood samples will be collected monthly (+/- 2 weeks) during chemotherapy and at the end of treatment (4 +/- 2 weeks post chemo) for ctDNA analysis
<sup>b</sup>Surveillance CT imaging will be performed at every 6 months (+/- 1 month) for the first 2 years and at the end of year 3

## Sample Size

A total sample size of 450 patients, with a 2:1 randomisation ratio, provides 80% power to demonstrate non-inferiority between arms A and B, assuming a type-I error rate α =0.05. The sample size calculations are based on one-sided Z tests, and 10% loss to follow-up rate. Assuming at least 10% of patients will have ctDNA detectable, a minimum of 30 patients with detectable post-operative ctDNA are expected to be enrolled into the ctDNA-informed treatment arm (Arm A). The patients in the control arm (Arm B) will be treated at clinician discretion according to standard practice, where based on previous studies the use of adjuvant chemotherapy would be expected to be 30% (45 patients).

The study was also powered to demonstrate statistical significance in the two key secondary endpoints:

- **TTR:** The study will achieve a 71% power to detect a non-inferiority margin difference of 5% in terms of 2-year TTR rate between Arms A and B. We assumed a 2-year TTR rate of 8% in Arm B. The test statistic used is the one-sided Z test (unpooled) with a targeted significance level 0.10.
- **Proportion of patients treated with adjuvant chemotherapy:** The study sample will achieve more than 99% power to detect a 20% absolute reduction of the proportion of patients treated with adjuvant chemotherapy in Arm A. The proportion of patients treated with chemotherapy in Arm B is expected to be 30%. The test statistic used is the two-sided Z test with unpooled variance. The significance level of the test was targeted at 0.05.

## General Considerations

### Timing of Analyses

The final analysis will be performed when all subjects have completed the 24-month follow-up visit or dropped out prior to the 24-month follow-up visit.

The final analysis will be performed on data transferred to the file DYNAMIC Data_For Analysis, having been documented as meeting the cleaning requirements of SOP_DYNAMIC Data Management, and after the finalisation and approval of this SAP document.

### Analysis Population

#### Full Analysis Population

- All eligible subjects who were randomised and had both post-op blood sample collected

#### Per Protocol Population

- All subjects who:
  - completed at least the 24-month surveillance CT scans, unless recurrence or death was documented prior
- Arm A (biomarker-guided) subjects who:
  - had successful ctDNA analysis performed
  - had positive ctDNA and received at least 12 weeks of chemotherapy
  - had negative ctDNA and received no more than 4 weeks of chemotherapy

The full analysis population will be used for primary efficacy analysis.

### Covariates and Subgroups

Important covariates that will be adjusted for in the primary analysis are:

- Primary tumour T stage (T3 vs T4)
- Lymph node yield (> 12 vs 12)
- Poor Differentiation (no vs yes)
- Lymphovascular invasion (no vs yes)
- Tumour perforation (no vs yes)
- Bowel obstruction (no vs yes)
- MMR status (deficient vs proficient)
- Centre (Metropolitan vs Regional)

Patients with proficient MMR tumours and any of the above clinico-pathologic risk features (T4, LN yield < 12, lymphovascular invasion, tumour perforation, bowel obstruction) will be classified as clinical high-risk; patients with deficient MMR tumours or none of the above risk features will be classified as clinical low-risk.

Exploratory subgroup-specific ***summary*** statistics will be presented as forest plot figures.

### Missing Data

The frequency of missing outcome (either binary or time to event) is expected to be minimal therefore no formal imputation method will be performed. However, any patient lost to follow-up within the first two years after randomisation without recurrence, will be coded 0 for the primary efficacy outcome and censored for the time to event outcomes.

For baseline variables, mean imputation method will be used to replace missing values when conducting covariate-adjusted analyses of the ctDNA-guided adjuvant treatment effect. Mean imputation is a reliable and straightforward approach to deal with missing baseline variables in clinical trials because the randomisation procedure is expected to guarantee that baseline variables are independent of treatment management strategy group (11). Mean values will be calculated from the non-missing values for the baseline variable using the pooled data from both groups. For categorical (coded 0, 1, 2) variables, the imputed mean will be rounded up or down to the nearest whole number (non-negative integer).

### Interim Analyses and Data Monitoring

#### Purpose of Interim Analyses

An interim analysis was conducted to demonstrate the feasibility of returning ctDNA results to clinician within the protocol specified timeframe. The interim analysis was planned after the first 20 patients have been randomized to ensure that the post-op ctDNA results can be fed back to the treating clinicians within 10 weeks after surgery.

#### Interim Meeting and Decision

A Data and Safety Monitoring Board (DSMB) was established to monitor study conduct and progress. The DSMB reviewed unblinded data to examine ctDNA turn-around time and treatment compliance once the first 20 patients had completed 10 weeks post surgery. After the interim review, the DSMC made the following recommendation: “*ctDNA results turn-around time was within the expected timeframe, consequently the trial was recommended to continue*”.

### Multi-Centre Studies

It is planned that the data from participating centres in this protocol will be combined, so that an adequate number of patients will be available for analysis. Centres will be stratified by metropolitan vs regional centres to account for potential treatment effect.

### Multiple Testing

No formal multiplicity testing adjustment will be performed. However, the outcomes are clearly ranked by degree of importance (primary, key secondary and secondary), and a limited number of prespecified subgroup analyses will be conducted.

## Summary of Study Data

All continuous variables will be summarised using the following descriptive statistics: n (non-missing sample size), mean, standard deviation, median, maximum and minimum. The frequency and percentages (where the denominator consists of sample availability for each associated variable) of observed levels will be reported for all categorical measures. In general, all data will be listed, sorted by site, treatment and subject, and when appropriate by visit number within subject. All summary tables will be structured with a column for each treatment in the order (Control, Experimental) and will be annotated with the total population size relevant to that table/treatment, including any missing observations.

### Subject Disposition

CONSORT diagram: a trial profile will illustrate patients’ progression through the study from initial screening for eligibility to completion of the final primary outcome assessment (see Figure 2).

**Figure 2.**
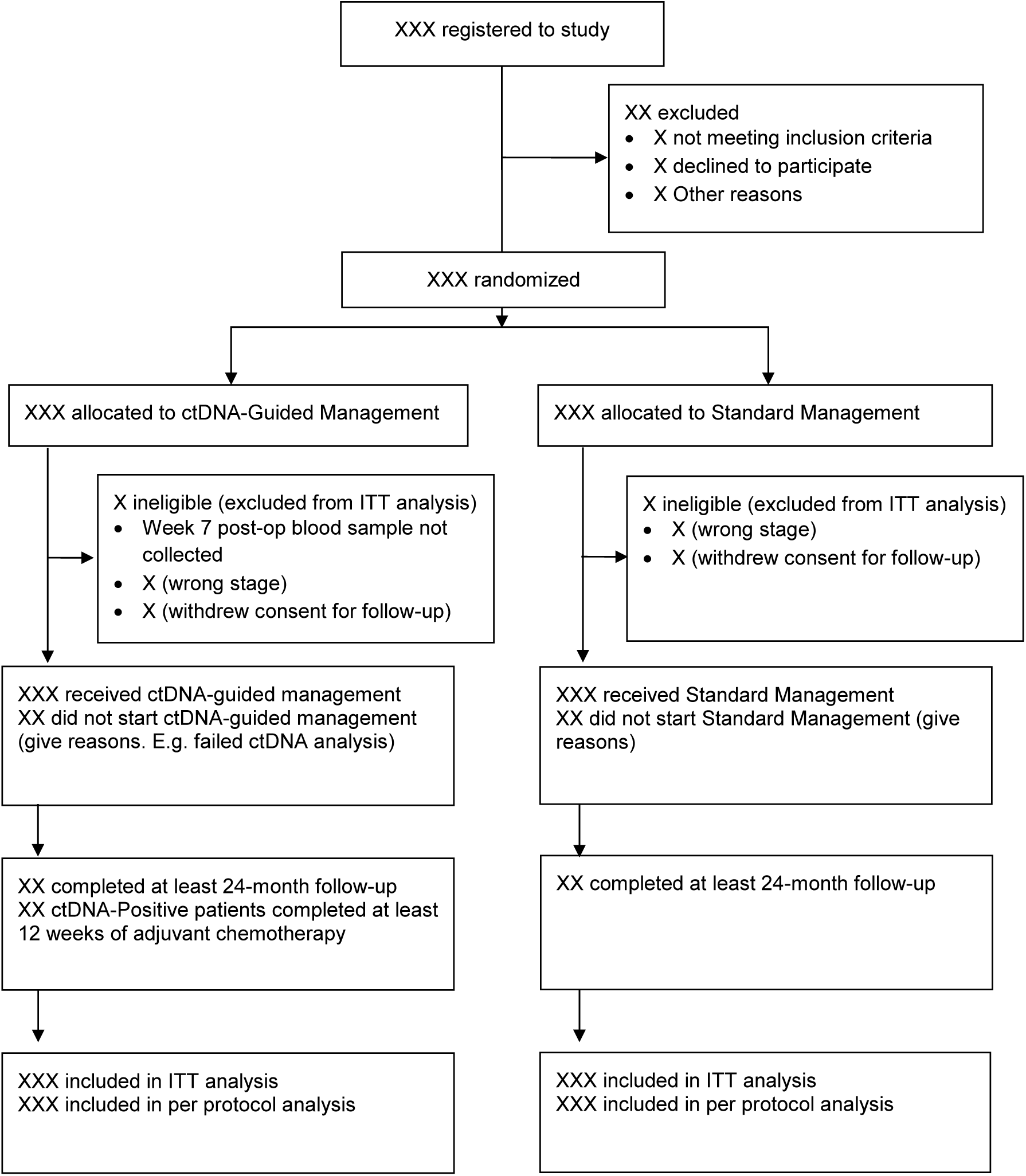
CONSORT Diagram.

### Protocol Deviations

Following randomisation, major deviations that would exclude a subject from the full analysis/ITT population include failure to obtain both post-op blood samples or failure to attend at least one follow-up assessment. Major deviations that would exclude a subject from the per protocol analysis include failure to complete the 24-month follow-up assessment, failure to complete at least 12 weeks of adjuvant chemotherapy for the biomarker-guided ctDNA positive patients, or treatment with at least 4 weeks of adjuvant chemotherapy for the biomarker-guided ctDNA negative patients.

### Demographic and Baseline Variables

Baseline characteristics, performance status, pathology details of the resected primary tumour, reasons associated with withdrawal and reason for non-compliance will be summarised by study arm using mean (sd) and/or median (range) for continuous variables and frequency (proportion) for categorical variables. Since any differences between randomised groups at baseline could only have occurred by chance, no formal significance testing will be conducted.

### Treatment Compliance

Treatment compliance will be assessed by duration of chemotherapy (treatment start and stop dates), reason for stopping chemotherapy and chemotherapy dose intensity (% of full dose received).

## Efficacy Analyses

Efficacy analyses will be conducted on the basis of intention to treat and will be summarised by treatment group. Efficacy analysis will be adjusted with the two stratification factors, type of participating centre (metropolitan vs regional) and T stage (T3 vs T4).

### General considerations

The analysis principles are as follows:

1. Efficacy analyses will be conducted both on an intention-to-treat (ITT) and per protocol basis (PP) but the ITT analysis remains the gold standard
2. All randomised patients will be analysed in the group to which they were assigned regardless of protocol violations. The only exception will be patients whose consent to use their data in the analysis is withheld or withdrawn.
3. All tests will be two-sided with a nominal level of alpha of 5%.
4. Effect of the treatment management strategy will be estimated as difference in proportions, means, odds ratio, or hazard ratio along with their 95% confidence interval (CI) and will be reported for all outcomes.
5. Adjusted analyses including the stratification factors will be performed as sensitivity analyses
6. Subgroup analyses will be carried out irrespective of whether there is a significant effect of treatment on the primary outcome.
7. P values will not be adjusted for multiplicity. However, the outcomes are clearly categorised by degree of importance (primary, key secondary and secondary), and a limited number of subgroup analyses are pre-specified.
8. All summary tables will be annotated with the total population size relevant to each treatment group.
9. P values ≥ 0.001 will be reported to three decimal places; P values less than 0.001 will be reported as ‘< 0.001’. The mean, SD and any other statistics other than quantiles will be reported to one decimal place greater than the original data. Quantiles, such as median, or minimum and maximum will use the same number of decimal places as the original data. Estimated parameters, not on the same scale as raw observations (e.g. regression coefficients), will be reported to three significant figures.
10. Analyses will be conducted primarily using SAS, version 9.4 or later and R 3.4.1 or later

### Primary Efficacy Analysis

The primary outcome analysis of a reduction in chemotherapy use without compromising the 2-year time-to-recurrence rate will be based on the intention-to-treat population. A sensitivity analysis will be performed using the per-protocol population. The per-protocol population is defined as those patients in Arm A with positive ctDNA who received at least 12 weeks of chemotherapy and Arm A ctDNA negative patients who received no more than 4 weeks of chemotherapy.

Non-inferiority of the primary outcome analysis will be tested by checking if the lower bound of the 95% confidence interval (CI) of the 2-year RFS rates difference between arms A and B exclude the non-inferiority margin of -8.5%. In particular, the proportion of patients alive without any signs or symptoms related to recurrence of colon cancer at 2 years follow-up along with the associated two-sided 95% CI for the difference between arms will be estimated. Non-inferiority of patient management informed by ctDNA (arm A) over standard management (arm B) will be accepted if the lower bound of the 95% CI around the estimated difference in the 2-year RFS rates lies above - 8.5%.

In addition, RFS will be analysed as time to event outcomes and will be described using the product limit method of Kaplan-Meier in each arm. Rates at 1-, 2-, and 3-years landmark points will be computed from the survival curves along with the associated 95% CI. This analysis will be conducted when the last patient reaches a minimum follow-up of 2 years. Additional analysis adjusting for non-compliance may be conducted if differing results for non-inferiority are observed between the intention-to-treat and per-protocol analysis.

### Secondary Efficacy Analysis

TTR will be analysed using similar methods as the primary outcomes. However the margin to declare non-inferiority will be 0.05 and one minus the Kaplan-Meier curve will be used to display the cumulative number of recurrence in either arm.

Overall survival will be described using the product limit method of Kaplan-Meier. Survival curve distribution differences between arms will be tested using log-rank test. Hazard ratios and corresponding 95% CI will be computed using the Cox proportional hazard model. The validity of the proportional hazards assumption will be tested using Shoenfeld residuals plots and corresponding test statistics. In contrast, if the proportional hazard assumption is violated (i.e. the hazard ratio is changing over time), an alternative robust measure named restricted mean survival time (RMST), which does not rely on a specific assumption, will be generated in each arm. Instead of HR then difference in RMST along with its 95% CI will be computed to evaluate the survival difference between groups.

The categorical secondary outcomes include reduction in chemotherapy use, and changes in ctDNA levels during adjuvant chemotherapy (the percentage of patients with initially detectable ctDNA where ctDNA remains detectable or becomes undetectable during chemotherapy) will be summarised by frequency and rate with its two-sided 95% Clopper-Pearson exact confidence interval. The denominator will be defined as the total number of patients for whom data is available. The difference between the two arms will be compared using a Pearson chi-squared test. In a case where expected count per cell is less than 5, Fisher’s exact test will be used. Overall treatment effect will be summarised using odds ratio and its 95% CI.

Association of changes in ctDNA level with risk of recurrence and time to recurrence will be analysed using logistic regression and Cox proportional hazard model respectively. All models will include randomization assignment as adjustment factor.

### Exploratory Efficacy Analyses

1. Subgroup: The primary endpoint and the key secondary endpoints will be assessed on a limited number of subgroups. The objective of this analysis is to assess potential differences in treatment strategies effects between subsets of patients, through an interaction term effect. Evidence of heterogeneity of treatment management effects among subgroups will be demonstrated by the level of statistical significance of the interaction term between treatment group and subgroup using either a logistic regression (for a binary outcome) or Cox regression (TTR, RFS and Overall survival) using a threshold of significance of 0.05. The subgroup analyses will remain exploratory and hypothesis-generating given the study is not specifically powered to test any treatment effect. Results will be presented as forest plots with P values for heterogeneity (interaction test) for each pair of subgroups displayed. The following factors will be investigated:
  a. Participating centre (metropolitan vs regional)
  b. T stage (T3 vs T4a vs T4b)
  c. Sex (male vs female)
  d. Age (> 70 vs 70 years)
  e. Primary tumour site (left vs right)
  f. Lymph Nodes Examined (> 12 vs 12)
  g. Tumour differentiation (poor vs well/moderate)
  h. Presence of lymphovascular invasion
  i. Bowel obstruction/perforation
  j. Mismatch repair status (MMR)
2. Association of ctDNA levels (mutant allele fraction) as a continuous variable, tumour mutation status and TIL status as a categorical variable with recurrence outcome will be explored.

### Safety Analyses

Given no treatments that are beyond standard of care were administered in this study, no safety analysis will be performed.

## Conclusion

We have developed a statistical analysis plan (SAP) for the DYNAMIC study. This plan will be followed to ensure high-quality standards of internal validity to minimise analysis bias. The SAP was approved and signed off by the study chairs and biostatistician on 28 Aug 2021.

## Data Availability

Data availability is not applicable for this manuscript

## List of Tables and Figures

A list of proposed tables and figures are displayed in this section.

## Table Listing

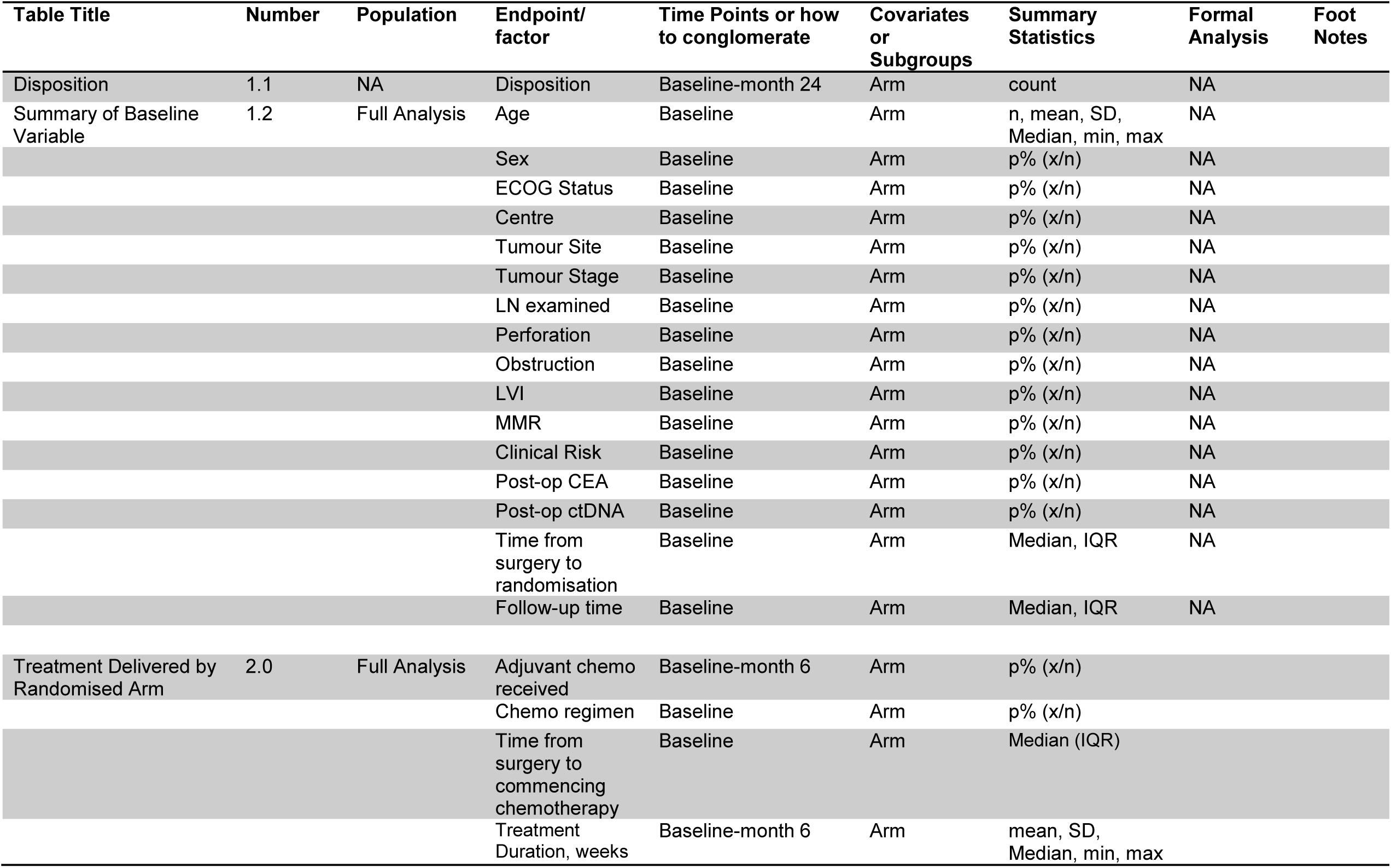
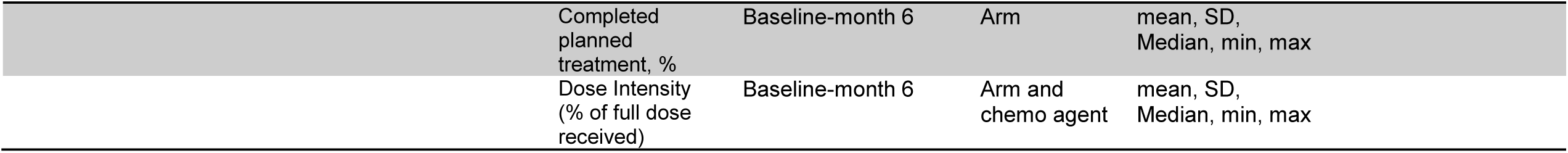

## Figure Listing

| Title | Number | Population | Type of graph | Horizontal Variables | Vertical Variables | Groupings | Statistics | Facets |
| --- | --- | --- | --- | --- | --- | --- | --- | --- |
| RFS | 1.1 | Full Analysis | KM | time | probability | Arm | Survival Estimates | NA |
| Difference of TTR/RFS/OS probabilities | 1.2 | Full Analysis | KM | time | Probability differences | Arm | Survival difference (95% CI) | NA |
| TTR | 1.3 | Full Analysis | KM | time | probability | Arm | Survival Estimates | NA |
| Overall Survival | 1.4 | Full Analysis | KM | time | probability | Arm | Survival Estimates | NA |
| Forest plot of RFS/TTR according to subgroup | 2.1 | Full Analysis | Forest Plot | HR | subgroup | Arm | Survival Estimates | NA |
| RFS/TTR by ctDNA status and management arm | 2.2 | Full Analysis | KM | time | probability | Arm and ctDNA status | Survival Estimates | NA |
| RFS/TTR by end of treatment ctDNA status | 3.1 | ctDNA-guided arm<br>ctDNA positive group – per protocol | KM | time | probability | End of treatment ctDNA status | Survival Estimates | NA |
| Difference of TTR/RFS probabilities | 3.2 | per protocol | KM | time | Probability differences | Arm | Survival difference (95% CI) | NA |
| Sankey plot of ctDNA clearance with adjuvant chemo and recurrence outcome | 3.3 | ctDNA-guided arm<br>ctDNA positive group – per protocol | Sankey plot | ctDNA timepoint and recurrence | ctDNA and recurrence status | ctDNA and recurrence status | Individual data |  |

## Notes

### Competing Interest Statement

BV is founder of, and hold equity in Thrive Earlier Detection and Personal Genome Diagnostics. KWK & NP are on the Board of Directors of, and consultants to, Thrive Earlier Detection. BV is consultant to and hold equity in NeoPhore. BV is a consultant to and holds equity in Catalio Capital Management. All other authors declare no competing interests.

### Clinical Trial

ACTRN12615000381583

### Funding Statement

This research is funded by: Australian National Health and Medical Research Council (NHMRC APP1085531); Medical Research Future Fund/NHMRC Investigators Grant (APP1194970)

### Author Declarations

Melbourne Health Human and Research Ethics Committee provided approval of the study protocol and patient consent form

